# Assessment of cross-reactive neutralizing antibodies induction against H5N1 clade 2.3.4.4b by prior seasonal influenza immunization in retail workers

**DOI:** 10.1101/2025.05.15.25327718

**Authors:** Andrea Arroyave, Henintsoa Rabezanahary, Aude Wantchecon, Vonintsoa Lalaina Rahajamanana, Ahmed Sahli, Mathieu Thériault, Denis Boudreau, Caroline Gilbert, Sylvie Trottier, Mariana Baz

## Abstract

Highly pathogenic avian influenza (HPAI) H5N1 has been a global concern since its emergence in 1997, causing widespread outbreaks in birds and sporadic human infections. The clade 2.3.4.4b H5N1 virus has rapidly expanded across continents, infecting numerous mammalian species. In 2024, it was detected in dairy cattle for the first time in the U.S., along with human cases following exposure. In Canada, the first human case of this avian influenza was reported in a critically ill adolescent in late 2024.

No human-to-human transmission has been documented, but concerns persist regarding mutations associated with enhanced virulence and human adaptation. Although seasonal influenza vaccines are not directed against H5N1, studies suggest that pre-existing immunity from prior infections or vaccinations may provide partial protection against severe H5N1 infections through cross-reactive immune response.

Given the ongoing circulation of avian influenza and the rise in human infections, this study evaluated the effectiveness of neutralizing antibodies developed against seasonal influenza viruses and their cross-reactivity with recent H5N1 strains. Serum samples from 194 retail sector workers in Quebec, collected between late 2021 and 2022, were analyzed using a microneutralization assay. While strong neutralizing activity was found against seasonal influenza viruses, no neutralizing antibodies were detected against H5N1 strains in either vaccinated or unvaccinated individuals. These findings emphasize the need to evaluate cross-reactive antibodies against the neuraminidase protein of H5N1, assess cellular immune responses potentially linked to protection against severe HPAI H5N1 infections and targeted vaccine strategies against recently emerged H5N1 influenza viruses.

## INTRODUCTION

Influenza viruses are highly variable RNA viruses responsible for annual epidemics in animals and humans, contributing to a significant global disease burden with up to 1 billion infections, 3–5 million cases of severe illness, and 650.000 deaths annually [1] as well as sporadic pandemics [2, 3]. Since its emergence in 1997, highly pathogenic avian influenza (HPAI) H5N1 has caused widespread outbreaks in birds, resulting in significant economic losses on farms and the death or culling of over 557 million birds globally between 2005 and 2023 [4, 5]. Sporadic human infections with H5N1 have also been reported following contact with infected animals, with over 950 documented cases worldwide and a fatality rate approaching 50% as of 2024 [6].

Following its initial detection, the H5N1 subtype evolved into multiple genetic clades due to the diversification of hemagglutinin (HA) and neuraminidase (NA) genes [7]. Among these, the clade 2.3.4.4b was first identified in domestic poultry in Asia in 2020, primarily affecting avian species [8]. However, this clade exhibited a remarkable ability for geographic expansion and host adaptation, leading to its spread across multiple continents. It was detected in wild birds in the United States in 2021 and subsequently spread rapidly across North America [9].

HPAI H5N1 clade 2.3.4.4b has raised global concern due to its rapid spread, severe impact, and capacity for interspecies transmission [10]. Spillover events have been reported in more than 50 species, raising concerns about its zoonotic potential [9, 11–13]. In early 2024, H5N1 clade 2.3.4.4b (genotype B3.13) was detected in dairy cattle in the U.S., marking the first known infections in this species, as well as human infections linked to exposure to infected poultry or dairy cattle, with clinical presentations ranging from conjunctivitis and mild respiratory symptoms to severe and fatal pneumonia and systemic complications [14–16].

In November 2024, Canada reported its first human case of H5N1 clade 2.3.4.4b (genotype D1.1) in a teenager from British Columbia who required intensive care [17]. A few months later, the first avian influenza-related fatality in the U.S. was recorded in Louisiana [18].

Due to the increasing frequency of spillovers and the enhanced zoonotic potential of clade 2.3.4.4b viruses [19], understanding population-level immunity is critical to assess pandemic risk and guide public health strategies [2, 20]. Although current seasonal influenza vaccines do not confer protection against avian influenza subtypes, they remain a cornerstone of prevention efforts [21]. These vaccines are strongly recommended to reduce the incidence of seasonal influenza infections and limit the risk of co-infections, which could potentially facilitate viral reassortment and consequently induce interhuman transmission or exacerbate disease severity [22, 23].

Recent studies have suggested that pre-existing immunity from prior infections or vaccinations against seasonal influenza may elicit cross-reactive immune responses, such as antibodies or memory T cells, that could modulate the clinical course of infection with emerging influenza strains [15, 24, 25]. This immune background may partially explain the mild symptoms observed in some reported H5N1 human infections [26–28].

However, the extent of cross-reactivity between seasonal immunity strains and circulating H5N1 variants remains unclear. Given its historically high pathogenicity and associated global burden [29], this poses a critical public health challenge, particularly in high-risk environments such as farms and live markets, where continued exposure may drive the emergence of new variants with increased zoonotic and pandemic potential [30, 31].

Therefore, this study aimed to evaluate whether pre-existing immunity from seasonal influenza infection and/or vaccination influences the immune response to currently circulating H5N1 viruses. We evaluated neutralizing antibody activity against five seasonal influenza strains and the cross-reactive response to two recently isolated H5N1 subtypes of avian and bovine origin, using serum samples collected from 194 retail workers in Quebec, Canada.

## METHODS

### 1. Study participants and samples

This study included 194 serum samples from a cohort of 304 participants derived from a previous study in retail workers in Québec City, Canada, which focused on the prospective evaluation of the immune response to SARS-CoV-2 in a high-risk occupational setting. Volunteer adults were recruited at the Centre Hospitalier Universitaire de Québec-Université Laval (CHUL) in Québec City between October 2021 and May 2022 [32]. All participants provided written informed consent, and information regarding participant characteristics, symptoms of respiratory infections, and vaccination status was collected (Table). This study was approved by the CHU de Québec-Université Laval Research Ethics Board (registration number 2021–5744), and all experiments were conducted following relevant guidelines and regulations.

|  | **Overall study population** |  |
| --- | --- | --- |
|  | **N = 194** | **%** |
| **Age (years), Mean±SD** | 44 ± 15.6 |  |
| **Age groups** |  |  |
| 18-59 | 157 | 80,9 |
| 60-75 | 37 | 19,1 |
| **Sex** |  |  |
| Female | 108 | 55,7 |
| Male | 86 | 44,3 |
| **Influenza vaccination** |  |  |
| Usually received | 37 | 19,1 |
| Received in the last year<sup>1</sup> | 30 | 15,5 |
| **Comorbidities** |  |  |
| Hypertension | 30 | 15,5 |
| Diabetes mellitus | 14 | 7,2 |
| Asthma | 16 | 8,3 |
| Chronic pulmonary disease | 5 | 2,6 |
| Cardiovascular disease | 6 | 3,1 |
| Liver disease | 2 | 1,0 |
| Cancer | 9 | 4,6 |
| Blood disorder | 1 | 0,5 |
| Immunosuppression | 7 | 3,6 |
| Chronic neurological disorder | 5 | 2,6 |
| Hypothyroidism | 10 | 5,2 |
| Other<sup>2</sup> | 48 | 24,7 |

Notes:
<sup>1</sup>Influenza vaccination 2021-2022 season
<sup>2</sup>Includes chronic inflammatory disease (arthritis, Crohn's and Raynaud's diseases, multiple sclerosis), allergies, and dyslipidemias.

### 2. Cells and viruses

The A/California/04/2009 (H1N1)pdm09, B/Phuket/3073/2013 (B/Yamagata lineage)-like, B/Austria/1359417/2021 (B/Victoria lineage)-like, A/Darwin/9/2021 (H3N2)-like, A/Victoria/4897/2022 (H1N1)pdm09-like influenza strains used in the serological assays were obtained from the National Institute for Biological Standards and Control (NIBSC) and propagated in Madin–Darby canine kidney cells overexpressing the α2,6 sialic acid receptor (ST6-GalI-MDCK cells), kindly provided by Y. Kawaoka from the University of Wisconsin, Madison, WI-USA [33], in Minimum Essential medium (MEM) supplemented with HEPES, 10mg/mL of puromycin and TPCK-treated trypsin (SAFC Biosciences, USA).

A/bovine/Texas/98638/2024 and A/British_Columbia/2032/2024 (H5N1) strains were obtained from the National Microbiology Laboratory (NML), Public Health Agency of Canada, and grown in Madin–Darby canine kidney cells (MDCK, ATCC CCL-34). Virus stocks were propagated and titrated in MDCK cells, then stored at −80 °C until use.

The studies involving seasonal influenza strains were conducted at Biosafety Level 2 (BSL-2), and those with the highly pathogenic H5N1 strains were carried out at Biosafety Level 3 (BSL-3) laboratories at the *CHU de Québec-Université Laval*.

### 3. Microneutralization assay

Live microneutralization (MN) assays to evaluate virus-neutralizing antibodies (NtAbs) were performed in MCDK cells, as described in previous studies [21]. In brief, serial two-fold dilutions of heat-inactivated serum samples (30 min at 56°C) were prepared starting from a 1:20 dilution. Equal volumes of serum and virus were mixed and incubated for 60 min at room temperature. The residual infectivity of the virus-serum mixture was then determined in MDCK cells, using four wells for each serum dilution. Positive and negative controls were included in each experiment. NtAb titer was defined as the reciprocal of the serum dilution that completely neutralized the infectivity of 100 TCID50 of each influenza virus strain, as determined by the absence of cytopathic effect on MDCK cells at day 4, as previously described [34, 35].

### 4. Statistical analysis

A titer of 10 was assigned to serum samples with undetectable (<20) NtAbs for mean titer calculations and statistical comparisons. Quantitative variables are described by their mean, standard deviation, and range. The significance of differences in NtAb titers among the 7 influenza virus strains was assessed using the Kruskal-Wallis one-way ANOVA, followed by Dunn’s multiple comparison test, with analysis performed in GraphPad Prism 10 (GraphPad Software, Inc., San Diego, CA). A p-value of < 0.05 was considered statistically significant.

## RESULTS

Neutralizing antibody titers against five seasonal influenza (H1N1, H3N2, B) and two H5N1 strains were evaluated in serum samples from study participants. Significant neutralizing responses were observed for seasonal influenza viruses, with NtAbs titers against A/California/04/2009 (H1N1)pdm09, A/Victoria/4897/2022 (H1N1), A/Darwin/9/2021 (H3N2), B/Phuket/3073/2013 (B/Yamagata) and B/Austria/1359417/2021 (B/Victoria) of 171 (range 10–8127), 16 (10–202), 81 (10–806), 32 (10–1810), and 12 (10–160), respectively. However, no measurable neutralizing activity was observed in the cohort against the H5N1 strains, A/bovine/Texas/98638/2024 and avian A/British_Columbia/2032/2024 (**Figure 1A**).

**Figure 1.**
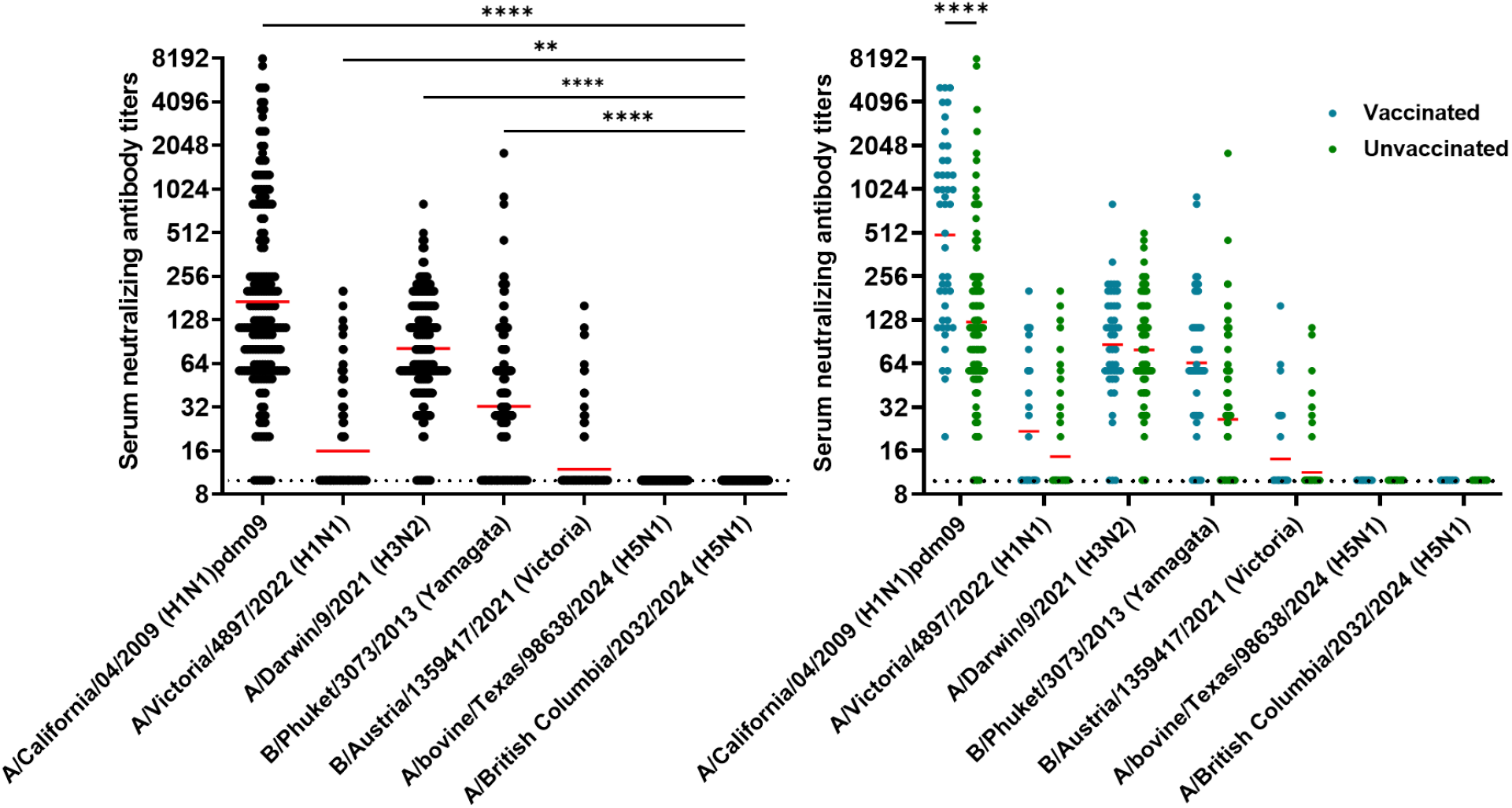
Serum neutralizing antibody titers against seasonal and avian influenza strains. **(A)** Serum NtAbs titers of all participants against influenza virus strains, including seasonal (H1N1, H3N2, and B) and avian (H5N1) strains. Each dot represents a single participant. Horizontal red bars indicate the median NtAbs titers of the group, with 95% confidence intervals (CI). The horizontal dashed line represents the lower limit of detection for the MN assay (neutralizing titer of 10). The samples that did not neutralize influenza viruses at a 1:20 serum dilution were assigned a neutralizing titer of 10 for graphic representation and statistical analysis. Statistical significance was assessed with Kruskal-Wallis one-way ANOVA, followed by Dunn’s multiple comparison test (\*\**p* ≤ 0.01, \*\*\*\**p* ≤ 0.0001). **(B)** Serum NtAbs titers by vaccination status. Blue dots represent vaccinated individuals, and green dots represent unvaccinated individuals. Red bars indicate median titers. Statistical significance is shown.

Participants were stratified by vaccination status. Although neither group showed detectable neutralizing activity against H5N1, vaccinated individuals, including those who regularly received influenza vaccination and those vaccinated during the 2021–2022 season, tended to have higher levels of NtAbs against seasonal influenza strains compared to unvaccinated individuals, with a significant difference observed for the A(H1N1)pdm09 virus (**Figure 1B**).

## DISCUSSION

In this study, we evaluated the NtAb responses against seasonal influenza viruses and the potential cross-reactive NtAb against recently emerged avian influenza viruses in the US and Canada in a unique cohort of participants. Significant neutralizing activity was observed against circulating seasonal influenza strains, with titers varying across subtypes. Notably, the influenza A(H1N1)pdm09 virus elicited the highest median NtAb titers, reflecting its continuous circulation and its inclusion in seasonal influenza vaccines since 2009.

Stratification by vaccination status revealed a tendency toward higher neutralizing antibody levels in vaccinated individuals compared to unvaccinated participants, with a statistically significant difference for A(H1N1)pdm09. These findings are consistent with previous studies demonstrating that prior influenza vaccination enhances immune responses against seasonal strains [36, 37].

Conversely, no measurable NtAbs against the A/bovine/Texas/98638/2024 and A/British_Columbia/2032/2024 H5N1 strains were detected, indicating a lack of cross-reactive humoral immunity to these HPAI viruses. This result suggests that existing immunity from other influenza viruses (like H1N1 or H3N2) does not offer significant protection against H5N1 infections through antibodies, and aligns with prior studies reporting limited or nonexistent serological protection against H5N1, with undetectable NtAb titers to the H5 HA protein in different populations [10, 38, 39]. While HA proteins of H5N1 strains have shown significant antigenic and genetic variability [40], the NA proteins of the A/H5N1 viruses of clade 2.3.4.4b, and those of A(H1N1)pdm09 viruses have showed 89.6% amino acid identity, suggesting thus that these NA proteins may have considerable conservation at some antigenic sites [10]. This raises the possibility that pre-existing immunity to H1N1 may confer partial protection via cross-reactive responses to conserved NA epitopes [26, 41].

While our results indicate limited humoral protection against H5N1, it is important to consider other immune mechanisms, such as cellular immunity, which may contribute to protection against severe disease [25]. T-cell responses induced by seasonal influenza vaccination have been shown to exhibit cross-reactivity with avian influenza strains, potentially modulating disease severity even in the absence of neutralizing antibodies [42]. However, the specific humoral and/or cellular immune responses contributing to a protective immune signature against emerging H5N1 strains remain to be fully defined.

In conclusion, our findings highlight the absence of cross-NtAb responses to two recently emerged H5N1 strains in a unique cohort, despite robust immunity to seasonal influenza. Although seasonal vaccination enhances protection against currently circulating strains, it appears insufficient to elicit cross-protective responses against antigenically distant viruses. These results underscore the need for the development of broadly protective influenza vaccines [43], and the continuous surveillance of antigenic evolution in circulating H5N1 viruses as a public health priority to anticipate and mitigate the risk of future avian influenza outbreaks [31, 44].

Importantly, licensed H5N1 vaccines are currently available for outbreak response, with reported seroconversion rates of 60-95% against circulating HPAI H5N1 clade 2.3.4.4b viruses [45]. However, their efficacy against emerging variants remains uncertain, and they may serve as bridging vaccines during early outbreak stages until strain-matched formulations are developed and deployed [46]. In this context, our findings reinforce the importance of proactively vaccinating high-risk groups, including individuals with occupational exposure to potentially infected animals and laboratory workers handling live H5N1 virus. These populations are key targets identified by the National Advisory Committee on Immunization, which recently recommended vaccination in elevated risk scenarios involving avian influenza exposure in Canada, aiming to prevent zoonotic infections and limit opportunities for viral adaptation and transmission [45].

## Data Availability

All data produced in the present study are available upon reasonable request to the corresponding author

## ETHICAL APPROVAL

This study was approved by the « Comité d’éthique de la recherche du CHU de Québec-Université Laval » (registration number 2021-5744). A unique, anonymized identifier was assigned to each participant and used to store the data and the samples.

## AUTHOR CONTRIBUTIONS

A.A, H.R. and M.B. designed the study. S.T. wrote the clinical protocol and recruited the participants. A.A, H.R, A.W., V.L.R., A.S. performed the experiments. M.B. supervised the study. M.T. performed data curation. A.A., H.R. and M.B. analyzed the data. A.A. wrote the manuscript. H.R., M.T., D.B., C.G., S.T., and M.B. revised the manuscript and provided scientific input. All authors have read and approved the final manuscript.

## ACKNOWLEDGEMENTS

The authors sincerely acknowledge the nurses and research coordinators at Centre de recherche du Centre Hospitalier Universitaire de Québec for their involvement in this study and all donors who agreed to participate in this research project. We are grateful to the National Microbiology Laboratory (NML), Public Health Agency of Canada, for providing H5N1 isolates used in this study.

